# Factors affecting antenatal corticosteroid use in low- and middle-income countries: facility characteristics, structural readiness, and past performance of CEmONC signal functions

**DOI:** 10.1101/2024.11.12.24317172

**Authors:** Wen-Chien Yang, Catherine Arsenault, Victoria Y. Fan, Nazia Binte Ali, Fadhlun Mohamed Alwy Al-beity, Emily R. Smith

## Abstract

**Background:** Antenatal corticosteroids (ACS) utilization is disproportionately limited in low- and middle-income countries where most global preterm newborns who could benefit from this intervention are born. Understanding the factors affecting ACS use is crucial for improving its uptake. This study aimed to investigate facility-level factors associated with ACS use in low-resource countries.

**Methods:** We used data from ten Service Provision Assessment surveys across nine countries. We restricted the sample to facilities that provided delivery services. Our primary outcome was recent ACS use, defined as having administered ACS within the past three months before the survey. We conducted mixed-effect log binomial regressions, with country as a fixed effect and sub-national regions as random intercepts, to explore the association between recent ACS use and facility characteristics, injectable corticosteroids and ultrasound availability, facility structural readiness, and past performance of nine Comprehensive Emergency Obstetric and Newborn Care (CEmONC) signal functions.

**Results:** This study included 6183 facilities from nine countries. Across eight countries with nationally representative data, only 22.7% (median, range 4.0% to 27.4%) of facilities that provided delivery services had used ACS recently. Urban facilities had a 21% higher likelihood of recent ACS use (95% CI 6%–38%) than rural facilities. Corticosteroid availability was associated with a 14% higher likelihood of recent ACS use (95% CI 1%–29%). Facilities in the highest readiness tertile were more likely to have recent ACS use than those in the lowest (RR 1.91, 95% CI 1.58–2.30). Each CEmONC signal function, except for assisted vaginal deliveries, was significantly associated with recent ACS use, with neonatal resuscitation having the largest effect (RR 2.62, 95% CI 1.93–3.55).

**Conclusion:** Facilities that had performed CEmONC services were more likely to administer ACS, highlighting the importance of provider knowledge, skills, and competence in managing obstetric and newborn emergencies for effective ACS provision.

## INTRODUCTION

Administering antenatal corticosteroids (ACS) to pregnant women with a high likelihood of preterm labor at gestational age (GA) 24 to 34 weeks is recommended by the World Health Organization (WHO) and major medical associations.(1–4) By accelerating fetal lung maturity, this intervention effectively reduces neonatal mortality and morbidity, particularly respiratory complications.(5) ACS use has been encouraged in low- and middle-income countries (LMICs), where 80% of preterm births occur globally.(6) However, its uptake remains limited in these countries. The WHO Born Too Soon report suggested prioritizing existing highly effective interventions, including ACS.(7) The International Federation of Gynecology and Obstetrics also listed ACS use among its PremPrep-5 initiative of effective and low-cost interventions to improve preterm outcomes.(8) Therefore, a thorough understanding of factors associated with low ACS use in LMICs is urgently needed.

Factors affecting ACS use in low-resource countries are complex.(9) These factors can be primarily grouped into several levels. At the national level, guidelines and policies on ACS use are fundamental. Previous studies have identified limited, unclear, or inconsistent national guidelines as major obstacles.(9–12) The 2023 WHO Model List of Essential Medicines includes dexamethasone, one of the two antenatal corticosteroids, under medicines for reproductive health and perinatal care.(13) A study assessing barriers to ACS use found that the absence of ACS from National Essential Medicines Lists was a significant barrier in half of the twelve African and Asian countries examined.(11) Another policy analysis for seven sub-Saharan countries showed that although most countries included ACS use in their national protocols, details on accurate GA assessment and criteria for identifying eligible women were often unclear.(12) At the facility level, poor healthcare quality—attributed to various factors, such as insufficient medicines and equipment for maternal and newborn care and lack of integration of ACS into regular perinatal care—hinders appropriate ACS use.(9, 11) At the individual level, providers’ knowledge gaps in ACS use and identifying preterm labor hinder its proper use.(9, 14, 15). Pregnant women, also at the individual level, may lack access to this intervention due to their underlying socioeconomic constraints; among those who have access, their perceptions of risks and benefits also affect ACS use.(9) Hence, untangling the complexity of factors affecting ACS use is critical for identifying priority actions.

Appropriate settings for safe and effective ACS use are challenging to define. The WHO recommends giving ACS when the following conditions are met: accurate GA assessment, a high likelihood of imminent preterm labor, no clinical maternal infections, and adequate childbirth and preterm newborn care available.(1) However, implementing some of these recommendations in low-resource settings poses practical challenges, raising discussions in the global community.(8, 16, 17) Facilities that can perform Comprehensive Emergency Obstetric and Newborn Care (CEmONC) are considered appropriate settings for administering ACS.(1, 11) Basic Emergency Obstetric and Newborn Care (BEmONC) includes a set of seven signal functions; CEmONC includes all BEmONC services and adds two additional functions: Cesarean section and blood transfusion.(18) This set of health services defines the capability of a facility to manage obstetric and newborn emergencies (19, 20), and health facilities in LMICs are designated as a BEmONC or CEmONC facility based on their capabilities.(18) It is important to note that many pregnant women who present with obstetric emergencies and require CEmONC services are likely to deliver preterm babies. While managing maternal emergencies, it is as crucial for providers to take a holistic approach and simultaneously implement interventions that improve preterm outcomes, such as administering antenatal corticosteroids. A study comparing indices to measure facility readiness noted that the past performance of CEmONC signal functions reflected a certain or minimally required level of inputs available and provider competence in managing obstetric emergencies.(21) However, no studies have examined the relationships between health facilities’ abilities to perform CEmONC signal functions and ACS utilization.

This study aimed to assess the complex relationships among ACS utilization, facility characteristics, corticosteroid and ultrasound availability, structural readiness, and the past performance of CEmONC signal functions.

## METHODS

### Study sample

We used Service Provision Assessment (SPA) health facility survey data from nine LMICs (ten surveys).(22) Our sample included three countries from Asia (Afghanistan 2018-2019, Bangladesh 2017-2018, Nepal 2021), one from the Caribbean (Haiti 2017-2018), and five from sub-Saharan Africa (the Democratic Republic of Congo (DRC) 2017-2018, Ethiopia 2021-2022, Malawi 2013-2014, Senegal 2018 and 2019, and Tanzania 2014-2015). We had previously conducted a study examining antenatal corticosteroid utilization in LMICs using the same data.(23) Most surveys employed a stratified random sampling strategy; some were national censuses. Unlike other surveys, the Afghanistan 2018-2019 survey primarily sampled urban hospitals. Since Senegal implemented continuous SPA over five consecutive years (24), we merged the 2018 and 2019 Senegal SPA data to achieve a comparable sample size to other surveys and to be consistent with our previous work.(23) The SPA questionnaire has five instruments, of which this study used data from the inventory instrument. We restricted our analyses to facilities that provided delivery services (normal delivery and/or Cesarean sections).

### Measures

The primary outcome was recent ACS use at the facility level, defined as having administered ACS within the past 3 months before the survey. Administering ACS within this timeframe was believed to reflect the facility’s functionality at the time of the survey.(18) We identified three main groups of facility-level factors to explore their relationships with recent ACS use. First, we included facility characteristics such as location (urban versus rural) and managing authority types. Because surveys used different groupings for managing authority types, we developed a standardized categorization with four groups (public, private for-profit, private not-for-profit/mission or faith-based, and others) **(Supplemental Table 1)**. Second, we identified 28 items essential for safe and effective ACS use based on the WHO recommendations, which could be grouped into four major categories: equipment, diagnostics, medicines and commodities, and guidelines (1); we used these items to develop facility structural readiness **(Supplemental Table 2)**. Based on the number of available items, we developed facility readiness tertiles that were country-specific to account for the vastly different contexts and survey years spanning from 2013 to 2021. A functional ultrasound was included separately because access to ultrasound in early pregnancy is crucial for accurate GA dating. The measure of a functional ultrasound was based on its availability in a health facility in general. The availability of at least one valid corticosteroid (injectable betamethasone or dexamethasone) was also included separately. However, we used its availability in the medicines for non-communicable diseases as a proxy because SPA does not survey corticosteroid availability in the maternal and newborn care section. We adopted the same approach as our previous study.(23) Additionally, we constructed several binary variables that indicated having at least one medical doctor, midwife, obstetrician/gynecologist, pediatrician, or specialist. Countries surveyed staff types in various ways of grouping. All countries surveyed the availability of medical doctors and midwives. Six countries (Afghanistan, Haiti, DRC, Malawi, Senegal, and Tanzania) assessed the availability of specialists in general but not specifically obstetricians/gynecologists or pediatricians. Conversely, three countries (Bangladesh, Nepal, and Ethiopia) collected data on the availability of obstetricians/gynecologists and pediatricians but not specialists. We used this data to construct a binary variable indicating at least one specialist. The development of staff variables is detailed in **Supplemental Table 3**. Lastly, we included each CEmONC signal function, defined as having ever performed each of them before the survey, in the model.

### Statistical analysis

In reporting descriptive statistics, we presented the proportion of facilities that had used ACS within the past three months before the survey by country and facility characteristics. We also reported the proportion of facilities that had performed each CEmONC signal function. In presenting outcome distribution across countries, we presented the median and range from eight countries, excluding Afghanistan, because of its different sampling strategy. To assess factors associated with recent ACS use, we conducted mixed-effect log binomial regressions to report adjusted relative risks (RR), including country as a fixed effect and sub-national regions within each country as random intercepts. We performed bivariate regressions that also included a fixed effect of country and random intercepts of sub-national regions. Modified Poisson regressions (with a robust standard error) were conducted to explore country-specific associations for each country.(25) This approach was chosen to obtain effect measures of relative risks and ensure model convergence with smaller sample sizes compared to the pooled regression models. We did not include sub-national divisions as random intercepts in within-country regressions. All analyses were performed using R.

## RESULTS

This study included 6,183 facilities of nine LMICs from ten SPA surveys. Across eight countries (excluding Afghanistan), about one-third of facilities (median 30.5%; range 14.6% to 89.6%) were in urban areas, whereas most (99.4%) facilities in Afghanistan were urban **(Table 1)**. Most facilities were public (median 83.6%; range 43.1% to 91.6%), followed by private for-profit facilities (median 9.8%; range 3.6% to 24.7%) and private not-for-profit/mission or faith-based facilities (median 8.7%; range 0.8% to 27.2%). Staffing of specialized professionals was generally suboptimal. Across eight countries, fewer than one-fifth or one-fifth of facilities had at least one medical doctor (median 17.3%; range 6.1% to 76.5%) or midwife (median 20.2%; range 7.7% to 95.9%). In Afghanistan, 39.6% of the facilities had at least one medical doctor, and most (93.4%) had at least one midwife. For the three countries that assessed the availability of obstetricians/gynecologists and pediatricians, fewer than 10% of the facilities had at least one obstetrician/gynecologist (median 8.9%; range 7.5% to 16.7%) and pediatrician (median 7.5%; range 5.1% to 12.5%). Similarly, only 9.1% (median; range 3.1% to 70.6%) of the facilities among the six countries that surveyed specialists had at least one specialist.

**Table 1.** Characteristics of surveys and facilities included in the analysis of 9 LMICs.

| Facility characteristics | Country and survey year |  |  |  |  |  |  |  |  |  |
| --- | --- | --- | --- | --- | --- | --- | --- | --- | --- | --- |
|  | Median (range)<sup>1</sup> | Afghanistan<br>2018-2019<br>(n=101) | Bangladesh<br>2017-2018<br>(n=822) | Nepal<br>2021<br>(n=807) | Haiti<br>2017-2018<br>(n=362) | DRC<br>2017-2018<br>(n=1354) | Ethiopia<br>2021-2022<br>(n=650) | Malawi<br>2013-2014<br>(n=542) | Senegal<br>2018& 2019<br>(n=593) | Tanzania<br>2014-2015<br>(n=952) |
| **Location** |  |  |  |  |  |  |  |  |  |  |
| Urban (%) | 30.5% (14.6% - 89.6%) | 100 (99.4%) | 326 (21.9%) | 485 (43.3%) | 145 (40.0%) | 293 (21.8%) | 391 (39.0%) | 82 (15.1%) | 542 (89.6%) | 278 (14.6%) |
| **Managing authority type (%)** |  |  |  |  |  |  |  |  |  |  |
| Public | 83.6% (43.1% - 91.6%) | 14 (8.2%) | 682 (83.6%) | 631 (91.6%) | 156 (43.1%) | 823 (61.9%) | 560 (91.3%) | 356 (65.5%) | 537 (85.3%) | 692 (83.6%) |
| Private for-profit | 9.8% (3.6% - 24.7%) | 71 (71.4%) | 104 (11.6%) | 154 (7.5%) | 89 (24.7%) | 154 (17.9%) | 75 (7.9%) | 25 (4.9%) | 37 (12.1%) | 70 (3.6%) |
| Private not-for-profit/mission or faith-based | 8.7% (0.8% - 27.2%) | 16 (20.4%) | 36 (4.9%) | 22 (0.8%) | 52 (14.3%) | 377 (20.2%) | 15 (0.9%) | 148 (27.2%) | 19 (2.6%) | 180 (12.5%) |
| Others | 0 % (0% - 17.9%) | 0 (0%) | 0 (0%) | 0 (0%) | 65 (17.9%) | 0 (0%) | 0 (0%) | 13 (2.5%) | 0 (0%) | 10 (0.3%) |
| **Staff (%)** |  |  |  |  |  |  |  |  |  |  |
| Has at least one medical doctor | 17.3% (6.1% - 76.5%) | 56 (39.6%) | 344 (27.1%) | 166 (7.1%) | 277 (76.5%) | 852 (31.1%) | 431 (23.2%) | 59 (10.8%) | 138 (11.3%) | 238 (6.1%) |
| Has at least one midwife | 20.2% (7.7% - 95.9%) | 97 (93.4%) | 121 (8.6%) | 176 (7.7%) | 277 (76.5%) | 233 (7.9%) | 252 (9.3%) | 520 (95.9%) | 461 (74.3%) | 577 (30.6%) |
| Has at least one obstetrician or gynecologist<sup>2</sup> | 8.9% (7.5% - 16.7%) | - | 213 (16.7%) | 196 (8.9%) | - | - | 191 (7.5%) | - | - | - |
| Has at least one pediatrician<sup>2</sup> | 7.5% (5.1% - 12.5%) | - | 164 (12.5%) | 168 (7.5%) | - | - | 144 (5.1%) | - | - | - |
| Has at least one specialist<sup>2</sup> | 9.1% (3.1% - 70.6%) | 86 (70.6%) | - | - | 165 (45.5%) | 107 (3.3%) | - | 31 (5.7%) | 89 (12.5%) | 133 (3.1%) |

<sup>1</sup> Median and range were obtained from eight countries excluding Afghanistan.
<sup>2</sup> Only Bangladesh 2017-2018, Nepal 2021, and Ethiopia 2021-2022 surveyed the availability of obstetricians or gynecologists, and pediatricians. Other countries surveyed the availability of specialists but did not survey specific types of staff.

### Recent ACS use

The proportions of facilities that had used ACS within the past 3 months varied, ranging from 4.0% in Tanzania to 27.4% in Ethiopia, with a median of 22.7% **(Table 2)**. Afghanistan had 59.4% of the facilities that used ACS recently. Recent ACS use also differed by facility characteristics **(Table 2)**. A larger proportion of urban facilities used ACS recently than rural facilities (44.5% versus 21.1%). Facilities with injectable corticosteroids and a functional ultrasound were more likely to use ACS in the past three months compared to those without. The proportion of facilities that had recently used ACS increased by readiness tertile: 14.6% for the lowest tertile, 21.9% for the middle, and 48% for the highest. Facilities with at least one medical doctor, midwife, or specialist were more likely to administer ACS recently than those without: 56.5% versus 13.2%, 38.8% versus 25.1%, 61.8% versus 23.4%, respectively. In addition, higher proportions of facilities that had provided any of the CEmONC services used ACS recently than those that had never performed the signal function.

**Table 2.** Mixed effect log binomial regression models of recent antenatal corticosteroid utilization.

|  | Proportion of facilities with recent ACS use (%)<sup>1</sup> | Bivariate regression<sup>2</sup> |  |  | Multivariate regression<sup>3</sup> |  |  |
| --- | --- | --- | --- | --- | --- | --- | --- |
|  |  | relative risk (RR) | 95% CI | p-value | Adjusted relative risk (aRR) | 95% CI | p-value |
| Country |  |  |  |  |  |  |  |
| Afghanistan 2018-2019 | 59.4% | 2.11 | 1.26 – 3.52 | **0.004** | 0.89 | 0.53 – 1.50 | 0.661 |
| Nepal 2021 | 8.2% | 0.45 | 0.28 – 0.71 | **0.001** | 0.34 | 0.22 – 0.51 | **<0.001** |
| Haiti 2017-2018 | 23.6% | 0.59 | 0.37 – 0.93 | **0.022** | 0.32 | 0.21 – 0.51 | **<0.001** |
| DRC 2017-2018 | 22.2% | 1.05 | 0.74 – 1.49 | 0.795 | 0.76 | 0.53 – 1.07 | 0.113 |
| Ethiopia 2021-2022 | 27.4% | 1.53 | 1.01 – 2.31 | **0.043** | 0.80 | 0.54 – 1.18 | 0.257 |
| Malawi 2013-2014 | 21.5% | 0.44 | 0.26 – 0.73 | **0.002** | 0.53 | 0.32 – 0.87 | **0.012** |
| Senegal 2018 and 2019 | 23.4% | 0.66 | 0.44 – 0.99 | **0.047** | 0.95 | 0.64 – 1.42 | 0.811 |
| Tanzania 2014-2015 | 4.0% | 0.35 | 0.24 – 0.51 | **<0.001** | 0.25 | 0.17 – 0.36 | **<0.001** |
| Bangladesh 2017-2018 | 23.2% | *ref* |  |  | *ref* |  |  |
| Facility characteristics |  |  |  |  |  |  |  |
| Location |  |  |  |  |  |  |  |
| Urban *versus* rural<sup>4</sup> | 44.5% *versus* 21.1% | 3.09 | 2.77 – 3.45 | **<0.001** | 1.21 | 1.06 – 1.38 | **0.004** |
| Managing authority type |  |  |  |  |  |  |  |
| Public | 28.0% | *ref* |  |  | *ref* |  |  |
| Private for-profit | 43.0% | 1.51 | 1.32 – 1.74 | **<0.001** | 0.78 | 0.66 – 0.91 | **0.002** |
| Private not-for-profit/faith or mission-based | 37.8% | 1.48 | 1.29 – 1.70 | **<0.001** | 0.85 | 0.73 – 0.99 | **0.039** |
| Others | 19.5% | 1.00 | 0.59 – 1.68 | 0.995 | 0.99 | 0.58 – 1.69 | 0.975 |
| Structural readiness |  |  |  |  |  |  |  |
| Corticosteroid availability *versus* unavailability<sup>5</sup> | 45.1% *versus* 18.9% | 2.72 | 2.43 – 3.04 | **<0.001** | 1.14 | 1.01 – 1.29 | **0.032** |
| Ultrasound availability *versus* unavailability<sup>5</sup> | 60.9% *versus* 19.3% | 5.00 | 4.48 – 5.57 | **<0.001** | 1.26 | 1.10 – 1.45 | **0.001** |
| Readiness tertile<sup>6</sup> |  |  |  |  |  |  |  |
| High | 48.0% | 7.52 | 6.37 – 8.89 | **<0.001** | 1.91 | 1.58 – 2.30 | **<0.001** |
| Middle | 21.9% | 2.27 | 1.91 – 2.70 | **<0.001** | 1.23 | 1.03 – 1.48 | **0.021** |
| Low | 14.6% | *ref* |  |  | *ref* |  |  |
| Staffing<sup>7</sup> |  |  |  |  |  |  |  |
| At least one medical doctor *versus* no medical doctor<sup>8</sup> | 56.5% *versus* 13.2% | 5.99 | 5.33 – 6.74 | **<0.001** | 1.43 | 1.22 – 1.67 | **<0.001** |
| At least one midwife *versus* no midwife<sup>8</sup> | 38.8% *versus* 25.1% | 3.17 | 2.84 – 3.55 | **<0.001** | 1.23 | 1.08 – 1.40 | **0.002** |
| At least one specialist *versus* no specialist<sup>8</sup> | 61.8% *versus* 23.4% | 4.37 | 3.91 – 4.90 | **<0.001** | 1.21 | 1.05 – 1.39 | **0.009** |
| CEmONC signal functions<sup>9</sup> |  |  |  |  |  |  |  |
| Ever provide parenteral antibiotics *versus* never | 37.8% *versus* 6.1% | 7.18 | 5.70 – 9.05 | **<0.001** | 1.48 | 1.15 – 1.90 | **0.002** |
| Ever provide parenteral oxytocin *versus* never | 33.1% *versus* 5.5% | 8.73 | 5.80 – 13.14 | **<0.001** | 1.77 | 1.15 – 2.73 | **0.010** |
| Ever provide parenteral anticonvulsants *versus* never | 46.2% *versus* 11.5% | 5.40 | 4.75 – 6.14 | **<0.001** | 1.65 | 1.43 – 1.91 | **<0.001** |
| Ever perform assisted vaginal delivery *versus* never | 37.1% *versus* 12.6% | 3.44 | 2.91 – 4.06 | **<0.001** | 1.12 | 0.93 – 1.34 | 0.224 |
| Ever perform manual removal of placenta *versus* never | 38.2% *versus* 7.2% | 5.62 | 4.57 – 6.91 | **<0.001** | 1.98 | 1.58 – 2.47 | **<0.001** |
| Ever perform removal of retained products *versus* never | 39.8% *versus* 11.8% | 3.74 | 3.24 – 4.32 | **<0.001** | 1.39 | 1.19 – 1.63 | **<0.001** |
| Ever perform neonatal resuscitation *versus* never | 36.7% *versus* 4.4% | 9.46 | 7.06 – 12.66 | **<0.001** | 2.62 | 1.93 – 3.55 | **<0.001** |
| Ever perform Cesarean sections *versus* never | 63.5% *versus* 13.1% | 8.01 | 7.12 – 9.01 | **<0.001** | 1.85 | 1.53 – 2.24 | **<0.001** |
| Ever provide blood transfusion *versus* never | 63.8% *versus* 15.1% | 6.69 | 5.99 – 7.47 | **<0.001** | 1.37 | 1.16 – 1.63 | **<0.001** |

<sup>1</sup> Recent ACS use was defined as using ACS within the past 3 months. For each country, the proportions were calculated considering facility sampling weight.
<sup>2</sup> Risk ratios (RR) were derived from bivariate regressions that included each independent variable separately, country fixed effects, and sub-national divisions as random intercepts.
<sup>3</sup> Adjusted risk ratios (aRR) were derived from mixed effect log binomial regression model that included all independent variables, country fixed effects, and sub-national regions as random intercepts.
<sup>4</sup> Rural is the reference level.
<sup>5</sup> Unavailability is the reference level.
<sup>6</sup> Readiness tertiles refer to country-specific readiness tertiles, calculated by the available numbers of equipment, diagnostics, medicines and commodities, and guidelines.
<sup>7</sup> These binary variables indicate facilities having at least one medical doctor, midwife, or specialist, which were constructed based on the surveyed staff types for each SPA.
<sup>8</sup> No medical doctor, no midwife, or no specialist is the reference level.
<sup>9</sup> Facilities that never performed each CEmONC signal function were viewed as the reference level

### CEmONC signal functions past performance

One-fifth of the facilities (19.0%, median of eight countries) had ever performed all nine CEmONC signal functions, compared to two-fifths (40.6%) in Afghanistan. Across eight countries and among nine signal functions, Cesarean sections and the provision of blood transfusions were performed in the smallest proportion of facilities since only CEmONC facilities were designated for these two services. Among seven BEmONC signal functions, the provision of parenteral anticonvulsant was performed in the smallest proportion of facilities (median 37.6%), while oxytocin administration was performed in the largest proportion (median 94.3%) **(Figure 1)**. Ethiopia was the only country among the eight countries where more than 80% of the facilities had performed each BEmONC signal function.

**Figure 1.**
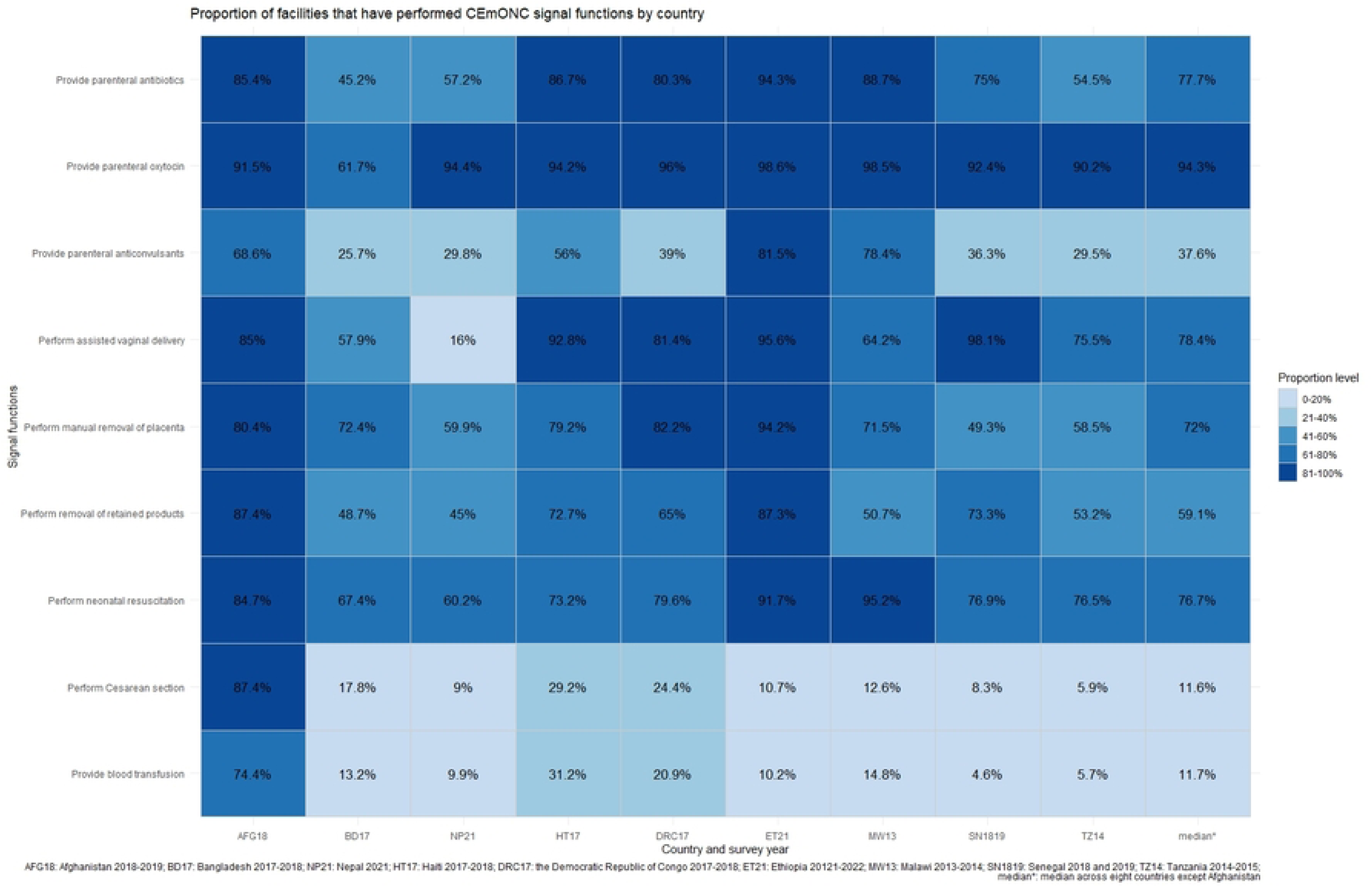
Proportion of facilities that have performed nine Comprehensive Emergency Obstetric and Newborn Care (CEmONC) signal functions

### Factors affecting recent ACS use

**Table 2** details the results of mixed-effect log binomial regressions, showing that facility characteristics, structural readiness, and past performance of CEmONC are associated with recent ACS use. Urban facilities had 21% higher recent ACS use than rural facilities (RR 1.21, 95% CI 1.06–1.39). Private for-profit facilities and private not-for-profit facilities, compared to public facilities, were associated with lower recent ACS use, although both types showed higher recent ACS use in bivariate regressions. The availability of injectable corticosteroids and ultrasound were associated with increased recent ACS use by 14% (RR 1.14, 95% CI 1.01–1.29) and 26% (RR 1.26, 95% CI 1.10–1.45), respectively. Facilities in the highest readiness tertile had 91% higher recent ACS use than those in the lowest (RR 1.91, 95% CI 1.58–2.30). Staffing types affected recent ACS use: facilities with at least one medical doctor, midwife, or specialist had increased recent ACS use by 43% (RR 1.43, 95% CI 1.22– 1.67), 23% (RR 1.23, 95% CI 1.08 to 1.40), and 21% (RR 1.21, 95%CI 1.05–1.39), respectively. Lastly, past performance of each CEmONC signal function, except for assisted vaginal deliveries, was significantly associated with more recent ACS use, with performing neonatal resuscitation having the largest effect (RR 2.62, 95%CI 1.93–3.56). Country-specific regressions were detailed in **Supplemental Table 4**.

## DISCUSSION

Our study showed the complex relationships between recent ACS utilization and facility-level factors. We found that recent ACS use was significantly higher in urban facilities, facilities with injectable corticosteroids and ultrasound available, facilities with the highest level of readiness, and facilities that had performed CEmONC services.

A consistent corticosteroid stock is fundamental for promoting ACS use.(26, 27) Our previous work found that only one-fourth of the facilities that provided antenatal care or delivery services in the same eight countries (excluding Afghanistan) had injectable corticosteroids available.(23) Interestingly, this study found a relatively small effect of corticosteroid availability (RR 1.14). As we used the availability of corticosteroids under the medicines for non-communicable diseases as a proxy, this approach might have overestimated their availability in maternal care. Thus, the effect size, while still statistically significant, is not as large as expected. The phenomenon might be explained by the idea that corticosteroid unavailability only partially contributed to the barrier to proper ACS use. In other words, while the availability of corticosteroids is essential, it does not guarantee their use. Our previous study also revealed the gap between corticosteroid availability and ACS use.(23) We suspect that the effect of corticosteroid availability might be offset by other factors, such as providers’ insufficient knowledge. Providers play a critical role in effective ACS utilization, including their awareness and knowledge of ACS, abilities to identify eligible women, and their perceived risks and benefits of ACS.(14, 15, 26, 28, 29) Similarly, the concern about the gap between availability and utilization also applies to facility readiness, meaning that high structural readiness does not always translate into good clinical practices; for example, the volume of delivery service of a facility is related to its provision of services for managing obstetric emergencies.(30) To triangulate factors affecting ACS use, by summarizing previous evidence, we proposed a framework for effective and safe ACS use at four levels **(Figure 2)**.

**Figure 2.**
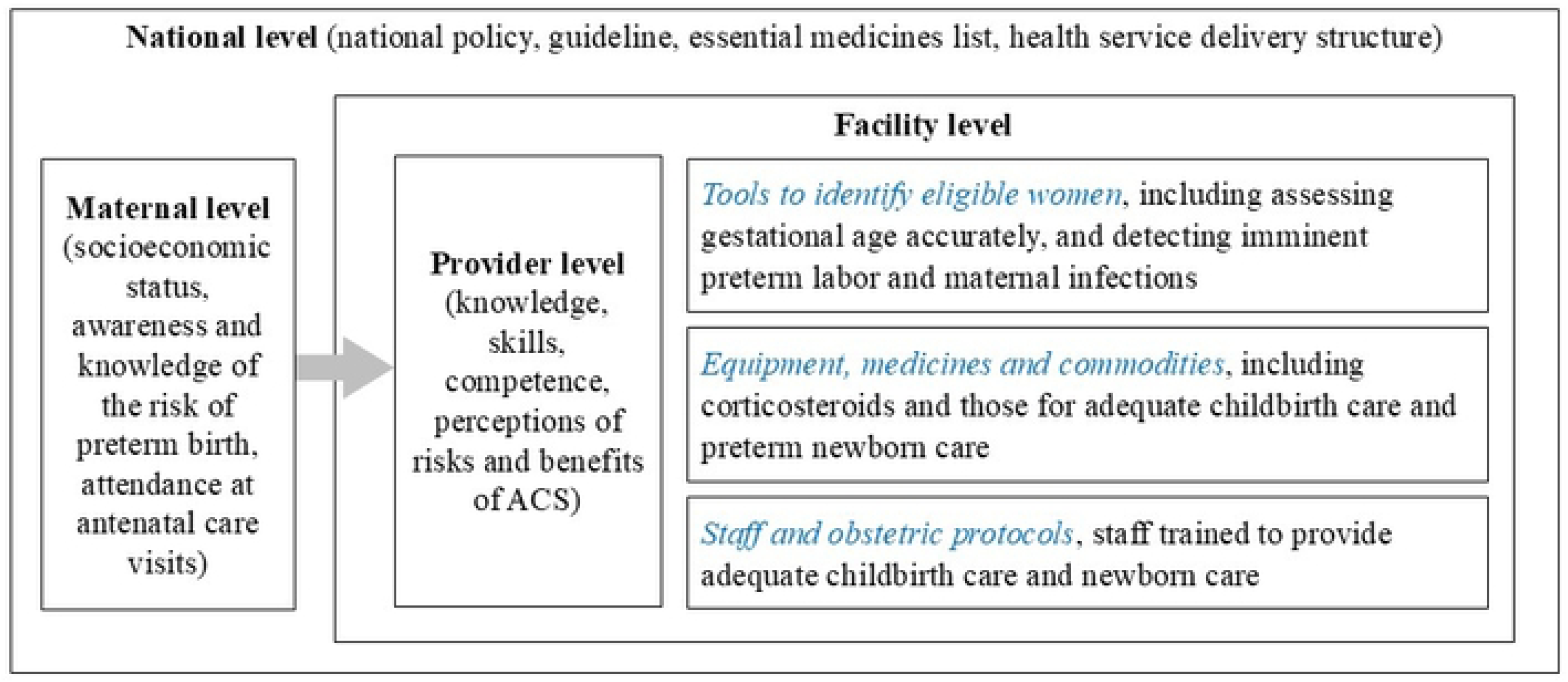
Framework for safe and effective antenatal corticosteroid utilization at four levels

While we found an association between ultrasound availability and recent ACS use, the effect size is relatively small, and the interpretation requires caution. First, ultrasound in early pregnancy, the gold standard for accurate GA dating, is uncommon in low-resource countries. Although its use is increasing in these countries (31), our previous work showed that only 7.3% of facilities that provided antenatal care or delivery services in eight LMICs had a functional ultrasound at the time of the survey.(23) A Bangladesh household survey of 700 women found that only 11% received an ultrasound examination in the first trimester.(32) Another critical point is that many women in LMICs initiate their antenatal care late (33, 34), resulting in missed opportunities. The prevalence of early antenatal care visits, defined as initiating before 14 weeks of gestation, was only 24.0% in LMICs versus 81.9% in high-income countries.(33) The limited ultrasound availability and late presentation to antenatal care might partly explain the small effect size in our findings. Additionally, the ultrasound measurement we used only reflected its availability at the time of the survey, which may not fully represent its availability in women’s first antenatal care visits, whether in the same facility or another.

Notably, this study identified strong associations between CEmONC signal functions and recent ACS use. There are a few potential explanations. First, CEmONC signal functions are meant to manage obstetric emergencies, which often involve a high likelihood of preterm labor. In these cases, providers should adopt a comprehensive approach to both save mothers’ lives and improve preterm newborn outcomes, including ACS administration to eligible women. For example, studies have demonstrated the benefits of administering ACS to women with severe preeclampsia.(35, 36) Second, this finding highlights the importance of provider knowledge, skills, and competence. The effect sizes for some CEmONC signal functions (RR 2.62 for neonatal resuscitation, RR 1.98 for performing manual removal of placenta, and 1.85 for performing Cesarean sections) are much larger than that for corticosteroid availability (RR 1.14). In other words, by controlling for all other factors, facilities with providers who have performed neonatal resuscitations are much more likely to administer ACS. This finding suggests that past performance of CEmONC signal functions reflects two critical aspects: first, providers’ knowledge, skills, and competence to perform the services, and second, the availability of a minimal level of medicines and equipment.(21) These two are crucial for delivering effective care; missing either will not work. Our findings have strong policy implications. Policymakers should prioritize provider training in emergent obstetric care while improving facility readiness and creating environments that encourage good clinical practice because in-service training alone does not deliver long-lasting effects.(37)

Our study has a few limitations. First, the countries in our sample differed in many aspects.(23) For example, national guidelines about ACS use vary. Some countries, such as Tanzania and Ethiopia, recommend administering ACS at the primary level of care, followed by referrals.(38–40) We recommend careful interpretations in any attempt to compare across countries. Second, there is a temporal discrepancy between corticosteroid availability and recent ACS use, with the former measured at the time of the survey and the latter defined as having provided ACS in the past 3 months before the survey. In addition, ACS utilization in the past 3 months does not represent the appropriateness of ACS use, meaning that the correct doses are given to eligible pregnant women at the right times. Third, we are aware of the collinearity between some independent variables in the regressions. For instance, facilities in the highest readiness tertile are likely to have the equipment, medicines, or staff required for CEmONC signal functions.

This study confirmed the complexity of factors affecting ACS utilization in LMICs. While corticosteroid availability is fundamental, providers’ knowledge and skills – reflected by the past performance of CEmONC services in our findings–affect safe and effective ACS administration. Policymakers should examine factors contributing to low ACS use at various levels and identify strategies to promote the implementation of this life-saving intervention.

## Data Availability

The data are publicly available on the DHS SPA website upon request. (https://dhsprogram.com/Data/)

## Disclosure of relationships and activities

All authors have completed the ICMJE uniform disclosure form at www.icmje.org/coi_disclosure.pdf and declared no support from any organization for submitted work; no financial relationships with any organization that might have an interest in the submitted work in the previous three years; no other relationships or activities that could appear to have influenced the submitted work.

## Ethical approval

This study was determined by the George Washington University Committee on Human Research, Institutional Review Board to be research that is exempt from IRB review (IRB# NCR235271).

## Author contributions

WCY, CA, VYF, and ESR conceptualized and designed the study and proposed an analysis plan. WCY conducted the analysis. CA, VYF, NBA, FMAA, and ESR provided inputs to the analysis results and suggested additional analyses. NBA reviewed the statistical codes and provided feedback. WCY, CA, and ESR drafted the manuscript. NBA and FMAA provided insights based on their expertise about the context and suggested areas for discussion. All authors critically reviewed the manuscript, made edits, and approved the final version of the manuscript for submission.

## Supporting information

Supplementary file

